# Comparing Phoneme and Word Recognition Test Outcomes in Adult CI users: Data Analysis from the AuDieT Study

**DOI:** 10.1101/2024.03.28.24304843

**Authors:** Enrico Migliorini, Jan-Willem A. Wasmann, Nikki Philpott, Bastiaan van Dijk, Birgit Philips, Wendy Huinck

## Abstract

**Purpose:** Current clinical measures used in cochlear implantation (CI) provide a broader view of speech recognition ability at word-level, often missing granular details contained at phoneme-level that may be valuable for CI mapping. This study evaluates how outcomes of Phoneme Recognition in Quiet tests (PRQ) differ from those of more commonly used word recognition tests (CVC) and outlines how these tests may be useful for different purposes in clinical adult CI care.

**Methods:** As part of the AuDiET (Auditory Diagnostics and Error-based Treatment) study, 23 adult postlingually deafened unilateral CI users underwent a battery of tests, including both PRQ and CVC tests. Their results were compared at the phoneme level, including an evaluation of fitness and error dispersion.

**Results:** PRQ had a significantly lower accuracy and fitness than CVC. The error patterns also tended to be less random and more systematic. Fitness correlated strongly and positively with accuracy, while error dispersion negatively correlated with accuracy.

**Conclusion:** There are clear differences between PRQ and CVC outcomes in absolute accuracy and error distribution. Comparing these tests might provide clinicians with more granular insights into which areas/phonemes to target during mapping, to achieve optimal speech recognition.

## Introduction

### Background

The cochlear implant (CI) is a highly successful sensory neuro-prosthesis. Through electrical stimulation of the auditory nerve, a CI can partially restore hearing in people with severe to profound sensorineural hearing loss. Since its invention, it is estimated that more than a million hearing-impaired people worldwide have received a CI (Zeng, 2022). Adult CI users score an average of 70%-80% speech recognition in quiet when using the latest implants, sound processors, and coding strategies (Zeng, 2022).

While this is an impressive result, some issues related to the post-implantation journey of CI users still need addressing. One of these is unexpectedly poor outcomes: cases in which a CI user achieves a much lower level than expected (Pisoni et al., 2017). There are multiple possible explanations for this variability: many of the factors which contribute to hearing, such as neural health in the spiral ganglion, are hard to measure precisely (Walia et al., 2023), the impact of various factors on performance seems to be variable (Blamey et al., 2013), and clinicians tend to overestimate the post-implantation performance of CI recipients based on pre-implantation data (Philpott, Philips, Donders, et al., 2023).

Clinical research tends to focus on good or exceptional outcomes (Moberly et al., 2016), resulting in an under-representation of people with poorer outcomes. There is a general lack of understanding on how to address the issue of poor performance. This uncertainty affects the otherwise excellent cost-effectiveness of CIs (Bond et al., 2009), and uncertainty about the potential benefits may dissuade candidates from undergoing the implantation.

The issue of addressing CI users who show poor speech understanding is compounded by the lack of patient-specific, standardised treatment guidelines. Whether considering fitting (i.e., adjusting the parameters governing the electrical stimulation) or training (i.e., rehabilitative exercises aimed at improving speech recognition), clinical practices vary significantly across different clinics (Wathour et al., 2021). In clinical fitting procedures for the Cochlear Nucleus system, the two most important settings which are individually determined are T-levels (just audible electrical stimulation level) and C-levels (most comfortable electrical stimulation level). T-levels are determined using a threshold-seeking method, while C-levels are set based on a CI user’s loudness rating. Most parameters other than T- and C-level are set to “default” (Vaerenberg et al., 2014). Surveys such as the aforementioned Vaerenberg et al., 2014 show significant variability in CI programming practices among clinicians and CI centres. The lack of standardised, evidence-based guidelines can lead different clinicians to adopt different approaches, although steps have been taken towards developing the Living Guidelines: consensus-based guidelines aimed at encouraging good clinical practices in clinics across the world.

While objective measures show a correlation with subjective threshold and comfort levels (de Vos et al., 2018), the correlation is, in general, weak; this means that the objective measures cannot be used to accurately predict T- and C-levels. However, it has been shown that creating MAPs based on either electrically evoked Compound Action Potential (eCAP) or electrically evoked Stapedius Reflex Threshold (eSRT) can lead to equivalent performance compared to behavioural subjective fitting (Craddock et al., 2003).

In recent years, several studies have been conducted aiming to develop individualised fitting and training interventions that take into account each participant’s unique challenges. The investigation of both fitting and training is important, as they represent different and complementary parts of the post-implantation journey of a CI user. Most of the studies attempting to define a new fitting paradigm focus on using objective, non-language-related measures to determine fitting levels: such tests may include electrode discrimination, pitch ranking or spectrotemporal sensitivity tests (Grasmeder et al., 2019; Van Opstal & Noordanus, 2023; Warren & Atcherson, 2023). Such “bottom-up” approaches stem from the assumption that fittings that provide well-audible stimuli across the full spectrum of covered frequencies would result in the best overall performance. Other approaches focus on imaging techniques, aiming to fit on the base of anatomical features, electrode positioning, and distance from the modiolus in order to leverage the tonotopic organization of the cochlea to prevent frequency drift due to misalignment of the electrode with the spiral ganglion cells (Jiam et al., 2019; Kurz et al., 2023).

It is known that CI users react to different cues than people with typical hearing thresholds (Moberly et al., 2014), which implies that delivering electrical stimulation with the aim of reproducing the neural activity of a normal ear as accurately as possible (i.e., making it as faithful as possible to the actual sound) may be a suboptimal approach. Enhancing the cues that CI users rely on, instead, may lead to better speech understanding. However, there is a very limited number of studies concerning how analysing speech recognition outcomes may provide insight into CI fitting (Holmes et al., 2012; Wathour et al., 2023), several of which centre around the Fitting to Outcome eXperts (FOX; Otoconsult NV, Antwerp, Belgium) system, an Artificial Intelligence (AI)-based fitting tool. The generally positive outcome of papers relating to speech-based interventions suggests that there is merit to this approach: however, the details of the interventions’ implementations have not been published in their entirety due to both their commercial nature and, in the case of FOX, the intrinsically hard-to-understand nature of AI. Therefore, the question of how clinicians may draft interventions based on their patients’ speech recognition outcomes remains partially unanswered.

As training interventions are complementary to fitting ones, it is worth mentioning how the concept of individualised auditory training has also been investigated: Magits et al. (Magits et al., 2023) reported similar effectiveness for personalised and non-personalised training programs, and in their literature review, Philpott et al. (Philpott, Philips, Tromp, et al., 2023) found variable effectiveness for both types of programs, although the personalisation of the training tended to refer to adaptive difficulty more than to a focus on the individual difficulties of each CI user. In this case, as well as investigating personalized fitting, we believed it worthwhile to evaluate whether personalized training based on the speech recognition issues of each CI user may be impactful.

The Auditory Diagnostics and Error-based Treatment (AuDiET) study was conceived as an investigation of the feasibility and effectiveness of individualised fitting and training interventions based on phoneme-level information on the participants’ errors. Its goal was to provide details on how CI users experiencing different challenges in sound recognition, respond to interventions aimed at addressing those challenges. Should an investigation of errors in speech audiometry provide valuable resources upon which fitting and training interventions may be based, this could lead the way towards a new approach to post-implantation clinical care. It may provide opportunities where users may test their hearing performance on their own between visits (apps for remote testing with self-administered procedures are already available, with varying degrees of functionality and validity (Wasmann et al., 2024)) and detailed information on their errors could be relayed to their clinicians for use in their follow-up.

In this paper we investigate whether there are significant differences in errors and error patterns when comparing the results of phoneme tests and word tests (as described in the Methods section). Next, differences between these tests are analysed to find out which test is better suited for designing interventions. This will be done by analysing their correlation scores, highlighting differences in accuracy scores, and dispersion both within participants and between the two different tests. Potential explanations for the presented results will be considered in the Discussion section.

## Methods

### Study design

In the AuDiET study, each participant undergoes five clinical visits. During Visit 1, baseline data is collected; during Visit 2, a fitting intervention is administered to the participant; during Visit 3 (set 2 weeks after Visit 2) the effects of the fitting intervention are evaluated, and the participant is given a personalised training programme; during Visit 4 (set 4 weeks after Visit 3) the training intervention is evaluated, and the participant stops training; finally, during Visit 5 (set 4 weeks after Visit 4) the retention of any effects is evaluated.

The AuDiET study itself is structured as a pre-post comparison; the analysis presented in this paper, however, is limited to the pre-intervention data collected from the study population. The study population is comprised of 23 (27 were recruited, 4 of which dropped out of the study or were excluded due to unforeseen technical issues) native Dutch-speaking adult CI users with a post-lingual onset of hearing loss and unilaterally implanted with a Cochlear® Nucleus™ implant model. The details of the population are highlighted in Table 1. All participants had at least one year of CI experience. Participants with abnormally formed cochleae, severe pre-implantation ossification, severe cognitive disorders, intense facial nerve stimulation, unaddressed tip fold-over or more than 4 malfunctioning electrodes were ineligible for the study.

**Table 1:** Information on the study population. ‘S’ stands for ‘Subject’, ‘M’ for ’Male’, ‘F’ for ‘Female’

| Subject No. | Age at inclusion<br>(years range) | Gender | CI experience<br>(y) | Implant<br>type | Etiology |
| --- | --- | --- | --- | --- | --- |
| S01 | 61-65 | M | 5 | CI522 | Idiopathic |
| S02 | 71-75 | M | 7 | CI512 | Family history,<br>otosclerosis |
| S03<br>(excluded) | - | - | - |  |  |
| S04 | 66-70 | F | 4 | CI522 | Idiopathic |
| S05 | 66-70 | F | 4 | CI422 | Family history, idiopathic |
| S06 | 71-75 | M | 4 | CI522 | Idiopathic |
| S07 | 81-85 | M | 3 | CI532 | Otitis media |
| S08 | 86-90 | M | 5 | CI522 | Idiopathic |
| S09 | 76-80 | F | 6 | CI522 | Skull trauma |
| S10<br>(dropout) | - | - | - | - | - |
| S11 | 81-85 | M | 5 | CI512 | Family history,<br>schwannoma |
| S12 | 76-80 | M | 8 | CI422 | Idiopathic |
| S13 | 66-70 | M | 5 | CI522 | Auditory neuropathy |
| S14 | 36-40 | F | 8 | CI422 | Congenital |
| S15 | 66-70 | M | 3 | CI512 | Sudden deafness,<br>idiopathic |
| S16 | 76-80 | M | 12 | CI512 | Unknown |
| S17<br>(excluded) | - | - | - | - | - |
| S18 | 66-70 | F | 7 | CI532 | Family history |
| S19<br>(excluded) | - | - | - | - | - |
| S20 | 66-70 | F | 10 | CI422 | Family history, idiopathic |
| S21 | 56-60 | F | 12 | CI24RE | Meningitis |
| S22 | 51-55 | F | 18 | CI24RE | Vestibular schwannoma |
| S23 | 71-75 | M | 7 | CI512 | Skull trauma |
| S24 | 66-70 | F | 5 | CI532 | Congenital, rubella |
| S25 | 71-75 | F | 7 | CI522 | Radiotherapy for vestibular schwannoma |
| S26 | 66-70 | F | 4 | CI422 | DFNA-9 |
| S27 | 76-80 | M | 7 | CI522 | Sudden deafness, idiopathic |

The study was submitted to the ethical committee, where it was approved and assessed as not falling under the jurisdiction of the Medical Research Involving Human Subjects Act (WMO) (NL-number: NL80521.091.22). Recruitment and testing took place at Radboud university medical center between 2022 and 2023.

During Visit 1, each participant underwent a test battery aimed at collecting a detailed dataset of their hearing capabilities. This battery included aided Pure Tone Audiometry (PTA), Spectrotemporal Sensitivity Assessment (SSA, (Van Opstal & Noordanus, 2023)), Phoneme Recognition in Quiet (PRQ, detailed below), Consonant-Vowel-Consonant (CVC, (Bosman & Smoorenburg, 1995)), and Digits Triplet Test (Smits et al., 2013). All tests except Pure Tone Audiometry were streamed via Direct Audio Cable (De Graaff et al., 2016) at a level of 65 input-related dBA to a Nucleus™ 6 test processor loaded with the participant’s most used MAP; Pure Tone Audiometry was instead performed in free-field using the modified Hughson-Westlake staircase procedure and ensuring the blocking of the contralateral ear in the case of residual hearing. The computer used for running all tests except PTA was a Lenovo Thinkpad T440 (Lenovo, Hong Kong, Hong Kong), connected to a RME Fireface UC external sound card (RME, Germany) to ensure consistent audio levels. Calibration was performed by directly reading the DSP input levels with a tool provided by Cochlear Ltd. and comparing streamed levels vs levels as acquired with the processor placed on a mannequin in a calibrated free field in a sound room. PTA was performed according to clinical routine on a calibrated clinical audiometer in a sound booth or quiet consultation room.

The data collected from the SSA and the DTT is not analysed and presented in this paper, as the goal was to investigate differences between phoneme and word tests; the results of those two tests were found to not be sufficiently granular for the purposes of investigating phoneme recognition. In the PRQ test, participants listened to triphones of the form /hVt/ or /aCa/, where V represents vowels or dipthongues (ɑ, a, ɑu, ɛ, e, ɛi, ø, ɪ, i, ɔ, u, o, ʏ, œy, y) and C represents consonants (b, d, f, ɣ, h, j, k, l, m, n, p, r, s, t, v, w, z) in the Dutch language. The participants were instructed to indicate what triphone they heard from the closed set of all possible options (vowels and consonants tested separately). The full set was presented in random order, 8 times for each consonant and 6 times for each vowel. The test software was developed specifically for this study using Python 3.

In the CVC test, the participants heard 15 lists of 12 meaningful Dutch CVC words each, taken from the Nederlandse Vereniging Audiologie (NVA) word list (Bosman & Smoorenburg, 1992), and were required to type what was heard as a response. This was then automatically converted into a triplet of phonemes in order to investigate errors on a phonemic level rather than judging words only as ‘correct’ or ‘incorrect’. The software for the CVC test was developed by Cochlear Ltd. and was previously validated in a clinical study (de Graaff et al., 2018). For further information on the tests itself, refer to the protocol as registered on https://clinicaltrials.gov/study/NCT05307952.

Both tests produced data points in the same format: arrays of stimulus-response pairings where the stimulus is the phoneme being presented, and the response is the phoneme the participant reported hearing. These arrays made up the primary outcome of the visit. In order to extract human-readable information from these data points, several data transformation techniques were applied. These include the calculation of accuracy (defined as the percentage of correctly identified phonemes), fitness (also describable as weighted accuracy, described in detail below), and error dispersion (defined in information theory as “the effective number of error classes per stimulus token” (Van Son, 1995)).

The measure of fitness was defined as 1 *−d* ( *stim , resp*) where ‘stim’ is the presented phoneme and ‘resp’ is the response. The distance function *d* is defined as the perceptual distance between the stimulus and the response, in such a way that if the stimulus and the response share some phonetic features, they are marked as being closer than ones that differ completely. The features of phonemes are those defined by the International Phonetic Association: voicing, place and manner for consonants, openness, place and rounding for vowels (International Phonetic Association, 1999). For instance, the distance between /p/ and /t/ can be set as 0.33 as they are both unvoiced plosives which only differ in place, while the distance between /p/ and /z/ is the maximum of 1, as they differ in voicing, place, and manner. In simple terms, fitness is lower when phonemes that are very different from each other are confused. The details of the implementation can be found in the supplemental materials. It is worth noting that, by definition, fitness dominates accuracy, i.e., for any given speech test, the fitness will always be higher than the accuracy. This is because every error is marked as a zero when calculating accuracy, while partial scores are possible when calculating fitness.

The data analysis aimed to compare the PRQ and CVC data, looking at correlations and differences between phoneme and word tests. This was done first by calculating the correlation coefficients between accuracy, fitness, and error dispersion in PRQ and CVC for each participant. The goal of this was to highlight how accuracy, fitness, and error dispersion scores can be useful for describing individual error patterns.

In order to assess within-visit test-retest reliability, random sampling was performed, splitting the data for each visit in half and checking for a significant change in accuracy. The test was repeated ten times and the results were averaged to reduce the chance of randomly selecting a split and returning a spurious result.

Next, significant differences between the scores’ distributions were investigated using the Wilcoxon signed rank test for paired samples. Finally, the correlation between accuracy in vowels and consonants was calculated for both PRQ and CVC. Results across the test are considered significant if their p-values are lower than 0.05 after Bonferroni-Holm correction (performed using (Gaetano, 2013)). All reported p-values have been adjusted in this way.

## Results

### Overview and normality test

The means, medians, and standard deviations of the computed measures can be found in Table 2. Using the Shapiro-Wilk test for normality, we found that the assumption of normality did not hold for fitness in vowels, either for PRQ or CVC tests, and for the error dispersion of consonants in CVC tests. For this reason, the non-parametric Wilcoxon test for paired samples was used instead of a parametric one.

**Table 2:** Aggregate measures and results of the Shapiro-Wilk test for each computed variable. Values below the 0.05 significance threshold are marked with an asterisk (*)

| Variable | Mean | Median | Standard deviation | P-value |
| --- | --- | --- | --- | --- |
| Accuracy of PRQ vowels | 69.13 | 72.06 | 16.49 | 0.09 |
| Fitness of PRQ vowels | 91.75 | 93.43 | 5.43 | **0.02\*** |
| Error dispersion of PRQ vowels | 0.68 | 0.58 | 0.35 | 0.20 |
| Accuracy of CVC vowels | 82.49 | 86.78 | 13.45 | 0.07 |
| Fitness of CVC vowels | 95.84 | 96.80 | 3.90 | **<0.01\*** |
| Error dispersion of CVC vowels | 1.09 | 1.10 | 0.70 | 0.19 |
| Accuracy of PRQ consonants | 66.62 | 69.12 | 19.45 | 0.33 |
| Fitness of PRQ consonants | 82.43 | 84.56 | 12.18 | 0.15 |
| Error dispersion of PRQ consonants | 0.96 | 0.88 | 0.69 | **0.02\*** |
| Accuracy of CVC consonants | 78.56 | 76.21 | 13.51 | 0.06 |
| Fitness of CVC consonants | 89.09 | 88.85 | 7.12 | 0.11 |
| Error dispersion of CVC consonants | 1.57 | 1.71 | 0.76 | 0.68 |
| Accuracy of PRQ vowels and consonants | 67.59 | 71.00 | 16.81 | 0.35 |
| Fitness of PRQ vowels and consonants | 85.61 | 87.95 | 9.30 | 0.08 |
| Error dispersion of PRQ vowels and consonants | 0.84 | 0.76 | 0.51 | 0.08 |
| Accuracy of CVC vowels and consonants | 79.74 | 79.84 | 13.34 | 0.07 |
| Fitness of CVC vowels and consonants | 91.13 | 91.26 | 6.01 | 0.11 |
| Error dispersion of CVC vowels and consonants | 1.40 | 1.56 | 0.71 | 0.57 |

### Correlation analysis – PRQ versus CVC scores

Error: Reference source not found shows the results of the CVC and PRQ tests for each subject. PRQ and CVC scores correlate strongly in all of accuracy (Pearson’s r: 0.90; *p*-value < 0.001), fitness (Pearson’s r: 0.87; *p*-value < 0.001) and error dispersion (Pearson’s r: 0.82; *p*-value < 0.001). These correlations remain present also when considering only vowels or only consonants.

CVC tests results have a significantly higher accuracy (Wilcoxon’s *p*-value < 0.001), fitness (Wilcoxon’s *p*-value < 0.001) and error dispersion (Wilcoxon’s *p*-value < 0.001) than PRQ test results. The average accuracy, fitness, and error dispersion of CVC tests were 80%, 91%, and 1.4, respectively; those of PRQ tests were 68%, 86%, and 0.84.

In order to ensure that this difference in error dispersion was not linked to the number of phonemes presented in CVC being higher than that of PRQ, random sampling was used to repeat the test 10 times using a randomly selected number of CVC phonemes, equal in number and in vowels/consonants percentage to the PRQ ones. The mean p-value of these ten randomly sampled tests was 0.044, which is higher (due to the much reduced sample size) but still significant.

The random sampling aimed at investigating test-retest reliability reported no significant change in the distribution of accuracy between any of the subsamples (the average *p*-value for a Wilcoxon test being 0.54).

Error: Reference source not found shows the data split between consonant and vowel scores. Significant correlations between the accuracies of the consonants vs vowels were found both in PRQ (Pearson’s r: 0.57; *p*-value: 0.002) and CVC (Pearson’s r: 0.95; *p*-value < 0.001).

### Correlation analysis – Fitness and Error Dispersion

Figure 3 shows that fitness correlated strongly and positively with accuracy (Pearson’s r: 0.97; *p*-value < 0.001).

**Figure 1:**
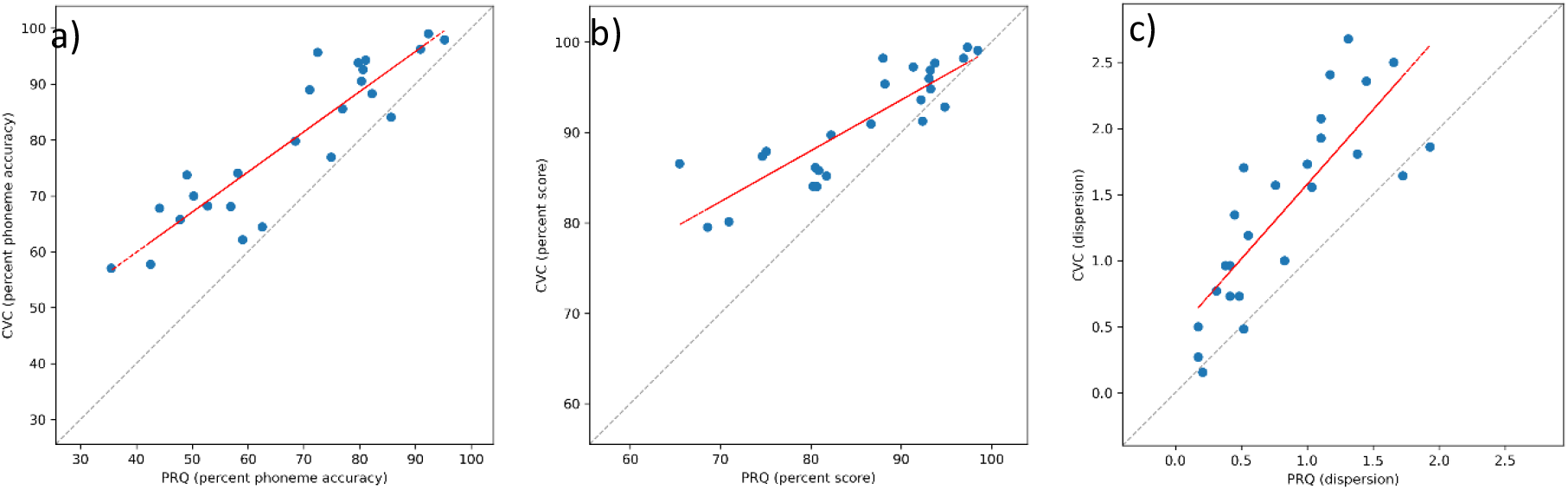
Distributions of PRQ and CVC scores for accuracy (a), fitness (b) and error dispersion (c). The red line indicates a linear regression through the data, the dotted grey line is the diagonal line where both scores are equal.

**Figure 2:**
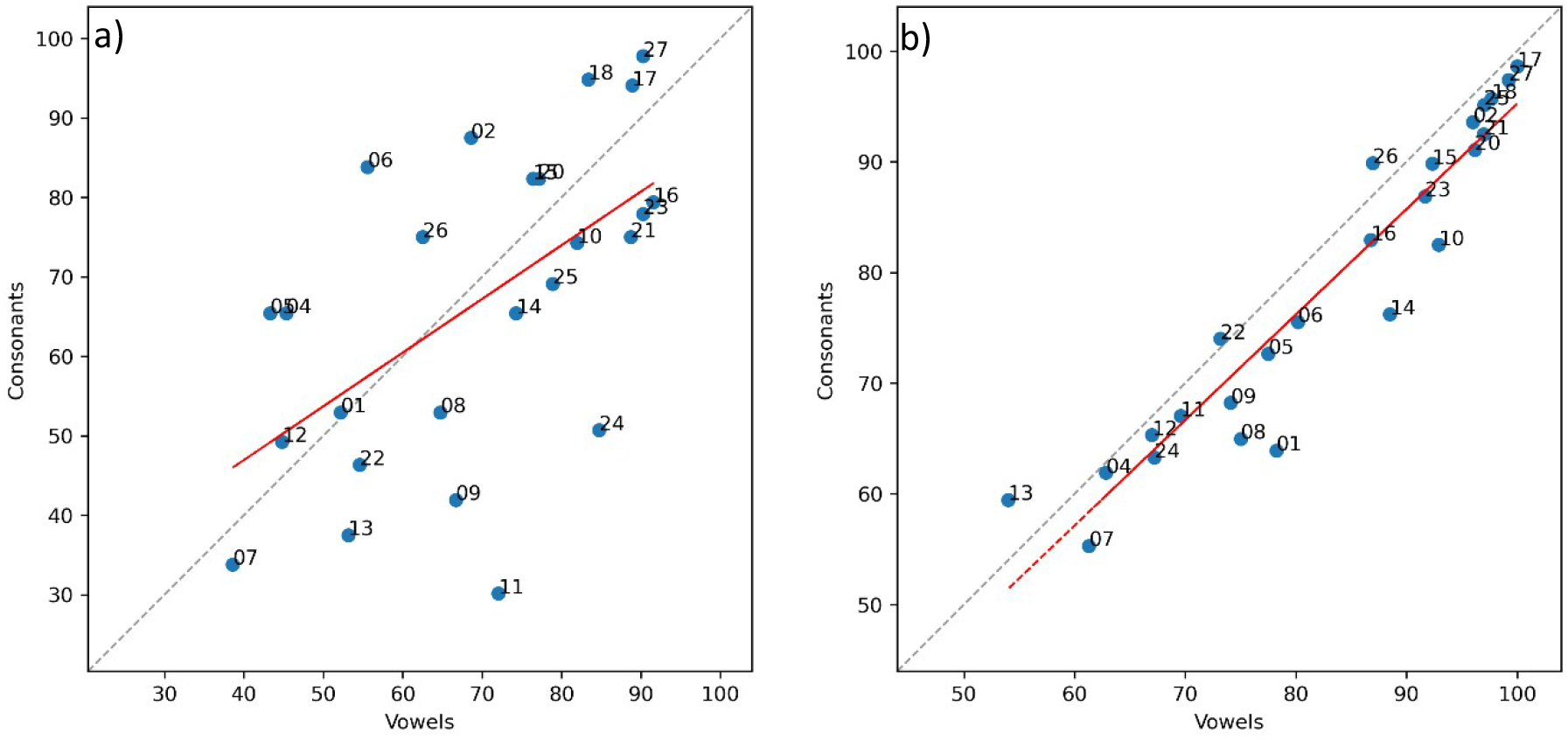
Distribution of accuracy in vowels and consonants for PRQ (a) and CVC (b) tests. The red line indicates a linear regression through the data, the dotted grey line is the diagonal line where both scores are equal. The numbers are Subject IDs.

**Figure 3:**
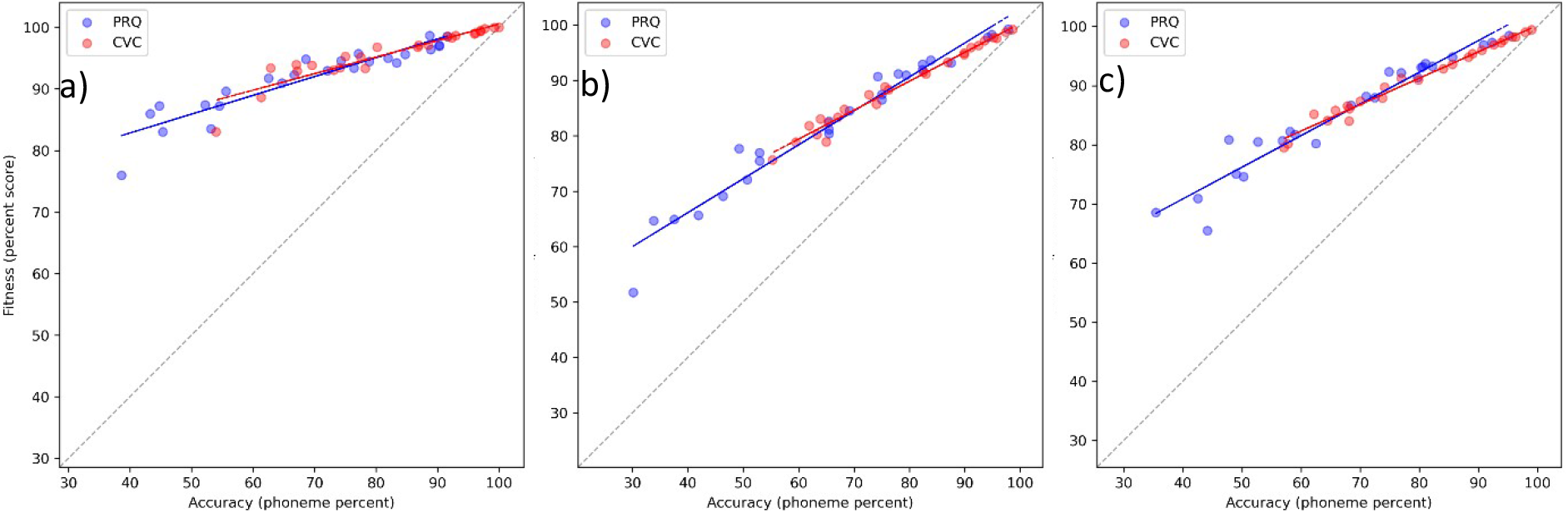
Distribution of fitness and accuracy for vowels (a), consonants (b) and overall (c). In these graphs the PRQ and CVC data is split by colour. The blue and red lines represent the linear regressions of PRQ and CVC data respectively. The dotted grey line is the diagonal line where both scores are equal.

Figure 4 shows that error dispersion correlates negatively with accuracy (Pearson’s r: -0.61; *p*-value < 0.001).

**Figure 4:**
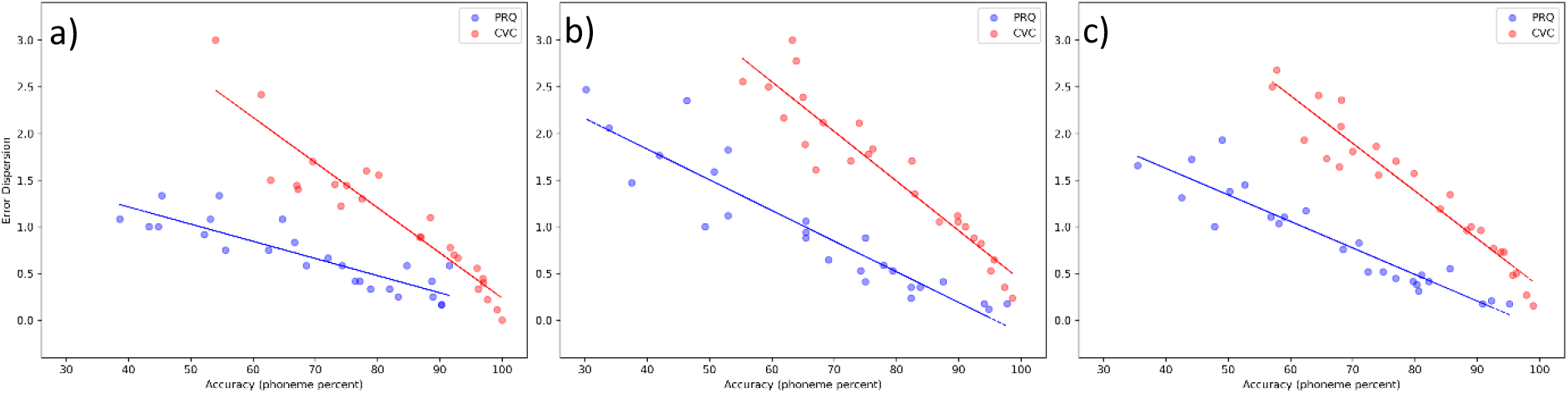
Distribution of error dispersion and accuracy for vowels (a), consonants (b) and overall (c). In these graphs the PRQ and CVC data is split by colour. The blue and red lines represent the linear regressions of PRQ and CVC data respectively. The dotted grey line is the diagonal line where both scores are equal.

### Power analysis

Using the G*Power software (Faul et al., 2007) to calculate the achieved power, it was found to be higher than 0.95 for each computer parameter, with the exception of the correlation between vowels and consonants for PRQ, where the achieved power was 0.91. This is usually considered to be a very high statistical power, resulting in a very low likelihood of a Type 1 error.

## Discussion

In this paper we have shown how, within the study participants, Phoneme Recognition in Quiet test scores differ from Consonant-Vowel-Consonants ones, having on average lower scores and more consistent error patterns (as shown by the lower error dispersion scores).

We have also shown that different measures, such as fitness and error dispersion, can provide additional information on a CI user’s speech recognition; we believe that there is a strong case to be made for the use of multiple scores (such as the aforementioned fitness and error dispersion) in clinical practice, delivering more granular insights to clinicians on their patients’ speech understanding. The strong correlation of fitness with accuracy is expected as the former dominates the latter and both measures are influenced by the number of errors made. However, in Figure 3, it is possible to see participants whose results are placed at a certain distance from the regression line, and for these comparing fitness and accuracy may be insightful. If the fitness of a participant’s PRQ test is close to the accuracy, it means that the participant is making large mistakes, confounding very different phonemes. A deeper analysis of their errors might then provide insight into their issues and, according to our hypothesis, make it so that an intervention could be designed to provide them with more easily recognisable cues. Conversely, a larger difference between fitness and accuracy means that the participant is confusing phonemes that are similar to each other, and they might be able to overcome this issue with practice if the differences between the cues of the confused phonemes are too small to intervene upon reliably. Furthermore, smaller errors (i.e., confusions between more similar phonemes) should be easier for a subject to correct when making use of contextual cues, such as in a meaningful sentence.

The negative correlation of error dispersion with accuracy can be explained by the fact that the fewer errors a CI user is making, the lower the probability will be that those errors distribute into a high number of categories. In a similar way to fitness, therefore, error dispersion is a measure that should not be interpreted by itself, but in relation to accuracy.

The three scores taken together can provide clinicians with more detailed information on their patients’ individual issues. A participant who, for example, has difficulties distinguishing similar phonemes such as /i/ and /e/ might be characterized by a fitness score significantly higher than accuracy (since the phonemes being mistaken are similar) and a low error dispersion score (since the errors are consistently made between those phonemes). Conversely, a participant who experiences difficulties with recognizing phonemic cues would show a higher error dispersion score; those issues might be addressed with training exercises aimed at practicing their phoneme discrimination and recognition capabilities (Philpott, Philips, Tromp, et al., 2023).

Considering the mechanics of speech comprehension can help interpret the differences between these tests. According to Erber’s hierarchy (Erber, 1977), there are four steps involved in listening: Detection, Discrimination, Recognition and Comprehension. By using meaningful words, CVC tests engage all skills involved in each of these steps, including cognition. Instead, PRQ tests, which use meaningless triphones, only involve the first three steps. CVC tests, by making use of cognition and top-down processes and being influenced by co-articulation, may mask certain auditory errors and cause others to appear. Subjects are told to expect meaningful words; therefore, they will correct perceptual errors, increasing the accuracy of the test. Sometimes, however, they might introduce non-perceptual errors by reporting a completely wrong, but meaningful word, e.g. (using English words for the sake of non-Dutch-speaking readers), mishearing the word ‘TOP’ as ‘TOG’ and reporting hearing ‘DOG’ instead.

The different correlations of words with consonants in PRQ and CVC could also be interpreted in a similar way. Considering that the CVC words are meaningful, correctly identifying the consonants in a word provides additional cues for identifying the vowels and vice versa. Conversely, in PRQ the participant can only rely on auditory cues. For instance, this could mean that a participant who perceives spectral components well but has issues with temporal ones might perform well in vowel recognition and more poorly in consonant recognition, which features temporal components more prominently.

These results would suggest that PRQ is an effective way to evaluate how a CI user experiences speech at a phonemic level, limiting the influence of co-articulation as well as cognition and comprehension skills. Arguably, reducing the effect of cognition might benefit clinicians aiming to adjust CI users’ fitting, as the test can help identify bottom-up errors at a perceptual level. These low-level errors are the ones which may be more effectively addressed by adjusting the CI user’s fitting. In contrast, CVC tests would be useful for replicating more closely speech in everyday life.

Another topic that may be further investigated is the implementation of phoneme tests at home. It would be little effort to implement the PRQ test on a mobile application and let CI users perform it at home; this would let clinicians have an overview of their patients’ speech recognition issues without devoting time to administering the test in a clinical environment. Research has shown that at-home tests have the potential to be as reliable as those run in a clinic (van Wieringen et al., 2021; Wasmann et al., 2024).

Finally, when considering how to assist a CI user optimally, the authors would recommend using a combination of accuracy, fitness, and error dispersion for both PRQ and CVC tests. This approach aims to paint a clearer picture of their individual difficulties in quiet. These scores might be further integrated by data-savvy clinicians with Confusion Matrices to pinpoint the phonemes that each participant has the most difficulties with.

### Limitations of the study

While PRQ appears to be a reliable test to evaluate CI users’ ability to identify phonemes without the additional variability introduced by co-articulation and cognition, it is worthwhile to take some time to discuss its limits. First, as the current study only included phonemes in quiet, results cannot be translated to speech recognition in noise. A follow-up to this study investigating whether these results hold for tests in noise is needed. Judging from previous studies (Goldsworthy et al., 2013), we can expect both PRQ and CVC scores to deteriorate with the introduction of noise; however, there would be merit in investigating which of the scores is most affected, and whether certain subsets of phonemes are more impacted by noise.

Second, the responses being presented as a multiple-choice test might introduce a form of McGurk effect (McGurk & MacDonald, 1976), inducing participants to report hearing phonemes that they read but did not hear. Similarly, they may develop a subconscious bias, repeatedly choosing one phoneme (for instance, the first one of the top row) when uncertain about what they heard.

Third, the test population consisted entirely of postlingually deafened, experienced CI users. A similar study using prelingually deafened or newly implanted CI users might show different results. The study by Magits et al. (Magits et al., 2023) included newly implanted subjects and found no differences between inexperienced and experienced users, so it may be worthwhile to check whether these results hold true for phoneme tests.

Finally, test-retest reliability needs further investigation in the context of multiple visits, and potentially in the context of self-administered tests over a long period.

## Conclusion

This paper presented Phoneme Recognition in Quiet testing as a valid integration to Consonant-Vowel-Consonant Speech Audiometry testing. We showcased how the two tests seem to measure different steps in the Erber hierarchy, respectively Recognition and Comprehension, and suggested that a framework based on the use of multiple measures (accuracy, fitness, and error dispersion) over both kinds of tests might provide audiologists with deeper insights into their patients’ unique and individual difficulties with speech recognition.

Further papers on the experimental parts of the AuDiET study will follow to investigate whether this data-driven, individualized approach to fitting and training can improve the post-implantation follow-up.

## Ethics Declarations

### Employment

Enrico Migliorini, Bastiaan van Dijk and Birgit Philips are employed by Cochlear Limited.

### Ethics approval

The study was carried on in conformity with the 1964 Declaration of Helsinki, the EU GDPR and all applicable guidelines for the Kingdom of the Netherlands and Radboud university medical center. The protocol was approved by Radboud university medical center’s METC.

### Informed Consent

All participants signed an informed consent module concerning their tests and gave explicit permission for their anonymous data to be shared.

## Data Availability

The data produced in the present study is not available at the moment.

## Supplemental Material

### Fitness calculation

Let *t* be a test made of *n* presentations of phonemes. Since *t* can be described as *n* couples of phonemes {*p*, *q*} where *p* is the presented phoneme and *q* is the given answer. The IPA features of *p* and *q* (voicing, place, and manner for consonants; rounding, place, and openness for consonants) are then compared. If *p* and *q* do not overlap in any of the features, the {*p*, *q*} couple is scored 0. If they overlap in one, the couple is scored 1/3; if they overlap in two, it is scored 2/3, and if they overlap completely then *p* = *q*, and the couple is scored 1. The average score over the *n* couples in the test *t* is the fitness of *t*.

## Bibliography

1. Blamey, P., Artieres, F., Başkent, D., Bergeron, F., Beynon, A., Burke, E., Dillier, N., Dowell, R., Fraysse, B., Gallégo, S., Govaerts, P. J., Green, K., Huber, A. M., Kleine-Punte, A., Maat, B., Marx, M., Mawman, D., Mosnier, I., O’Connor, A. F., … Lazard, D. S. (2013). Factors affecting auditory performance of postlinguistically deaf adults using cochlear implants: an update with 2251 patients. Audiology & Neuro-Otology, 18(1), 36–47. 10.1159/000343189

2. Bond, M., Mealing, S., Anderson, R., Elston, J., Weiner, G., Taylor, R. S., Hoyle, M., Liu, Z., Price, A., & Stein, K. (2009). The effectiveness and cost-effectiveness of cochlear implants for severe to profound deafness in children and adults: a systematic review and economic model. Health Technology Assessment (Winchester, England), 13(44). 10.3310/HTA13440

3. Bosman, A. J., & Smoorenburg, G. F. (1992). Woordenlijst voor spraakaudiometrie. Nederlandse Vereniging Voor Audiologie, Utrecht. https://scholar.google.com/scholar_lookup?title=Woordenlijst+Voor+Spraakaudiometrie&author=A.J.+Bosman&author=G.F.+Smoorenburg&publication_year=1992&

4. Bosman, A. J., & Smoorenburg, G. F. (1995). Intelligibility of Dutch CVC syllables and sentences for listeners with normal hearing and with three types of hearing impairment. Audiology : Official Organ of the International Society of Audiology, 34(5), 260–284. 10.3109/00206099509071918

5. Craddock, L., Cooper, H., van de Heyning, P., Vermeire, K., Davies, M., Patel, J., Cullington, H., Ricaud, R., Brunelli, T., Knight, M., Plant, K., Cafarelli Dees, D., & Murray, B. (2003). Comparison between NRT-based MAPs and behaviourally measured MAPs at different stimulation rates--a multicentre investigation. Cochlear Implants International, 4(4), 161–170. 10.1179/CIM.2003.4.4.161

6. de Graaff, F., Huysmans, E., Merkus, P., Theo Goverts, S., & Smits, C. (2018). Assessment of speech recognition abilities in quiet and in noise: a comparison between self-administered home testing and testing in the clinic for adult cochlear implant users. International Journal of Audiology, 57(11), 872–880. 10.1080/14992027.2018.1506168

7. De Graaff, F., Huysmans, E., Qazi, O. U. R., Vanpoucke, F. J., Merkus, P., Goverts, S. T., & Smits, C. (2016). The Development of Remote Speech Recognition Tests for Adult Cochlear Implant Users: The Effect of Presentation Mode of the Noise and a Reliable Method to Deliver Sound in Home Environments. Audiology & Neuro-Otology, 21 Suppl 1(1), 48–54. 10.1159/000448355

8. de Vos, J. J., Biesheuvel, J. D., Briaire, J. J., Boot, P. S., van Gendt, M. J., Dekkers, O. M., Fiocco, M., & Frijns, J. H. M. (2018). Use of Electrically Evoked Compound Action Potentials for Cochlear Implant Fitting: A Systematic Review. Ear and Hearing, 39(3), 401–411. 10.1097/AUD.0000000000000495

9. Erber, N. (1977). Evaluating speech-perception ability in hearing impaired children. In Childhood Deafness (pp. 173–181). Grune & Stratton.

10. Faul, F., Erdfelder, E., Lang, A. G., & Buchner, A. (2007). G*Power 3: a flexible statistical power analysis program for the social, behavioral, and biomedical sciences. Behavior Research Methods, 39(2), 175–191. 10.3758/BF03193146

11. Gaetano, J. (2013). Holm-Bonferroni sequential correction: an Excel Calculator.

12. Goldsworthy, R. L., Delhorne, L. A., Braida, L. D., & Reed, C. M. (2013). Psychoacoustic and Phoneme Identification Measures in Cochlear-Implant and Normal-Hearing Listeners. Trends in Amplification, 17(1), 27. 10.1177/1084713813477244

13. Grasmeder, M. L., Verschuur, C. A., van Besouw, R. M., Wheatley, A. M. H., & Newman, T. A. (2019). Measurement of pitch perception as a function of cochlear implant electrode and its effect on speech perception with different frequency allocations. International Journal of Audiology, 58(3), 158–166. 10.1080/14992027.2018.1516048

14. Holmes, A. E., Shrivastav, R., Krause, L., Siburt, H. W., & Schwartz, E. (2012). Speech based optimization of cochlear implants. International Journal of Audiology, 51(11), 806–816. 10.3109/14992027.2012.705899

15. International Phonetic Association. (1999). Handbook of the International Phonetic Association: A guide to the use of the International Phonetic Alphabet. Cambridge University Press.

16. Jiam, N. T., Gilbert, M., Cooke, D., Jiradejvong, P., Barrett, K., Caldwell, M., & Limb, C. J. (2019). Association Between Flat-Panel Computed Tomographic Imaging-Guided Place-Pitch Mapping and Speech and Pitch Perception in Cochlear Implant Users. JAMA Otolaryngology--Head & Neck Surgery, 145(2), 109–116. 10.1001/JAMAOTO.2018.3096

17. Kurz, A., Herrmann, D., Hagen, R., & Rak, K. (2023). Using Anatomy-Based Fitting to Reduce Frequency-to-Place Mismatch in Experienced Bilateral Cochlear Implant Users: A Promising Concept. Journal of Personalized Medicine, 13(7). 10.3390/JPM13071109

18. Magits, S., Boon, E., De Meyere, L., Dierckx, A., Vermaete, E., Francart, T., Verhaert, N., Wouters, J., & Van Wieringen, A. (2023). Comparing the Outcomes of a Personalized Versus Nonpersonalized Home-Based Auditory Training Program for Cochlear Implant Users. Ear and Hearing, 44(3), 477–493. 10.1097/AUD.0000000000001295

19. McGurk, H., & MacDonald, J. (1976). Hearing lips and seeing voices. Nature 1976 264:5588, 264(5588), 746–748. 10.1038/264746a0

20. Moberly, A. C., Bates, C., Harris, M. S., & Pisoni, D. B. (2016). The Enigma of Poor Performance by Adults with Cochlear Implants. Otology & Neurotology : Official Publication of the American Otological Society, American Neurotology Society [and] European Academy of Otology and Neurotology, 37(10), 1522. 10.1097/MAO.0000000000001211

21. Moberly, A. C., Lowenstein, J. H., Tarr, E., Caldwell-Tarr, A., Welling, D. B., Shahin, A. J., & Nittrouera, S. (2014). Do adults with cochlear implants rely on different acoustic cues for phoneme perception than adults with normal hearing? Journal of Speech, Language, and Hearing Research : JSLHR, 57(2), 566. 10.1044/2014_JSLHR-H-12-0323

22. Philpott, N., Philips, B., Donders, R., Mylanus, E., & Huinck, W. (2023). Variability in clinicians’ prediction accuracy for outcomes of adult cochlear implant users. International Journal of Audiology. 10.1080/14992027.2023.2256973

23. Philpott, N., Philips, B., Tromp, K., Kramer, S., Mylanus, E., & Huinck, W. (2023). Phoneme Training for Adult Cochlear Implant Users: A Review of the Literature and Study Protocol. Journal of Speech, Language, and Hearing Research : JSLHR, 66(12), 5071–5086. 10.1044/2023_JSLHR-23-00335

24. Pisoni, D. B., Kronenberger, W. G., Harris, M. S., & Moberly, A. C. (2017). Three challenges for future research on cochlear implants. World Journal of Otorhinolaryngology - Head and Neck Surgery, 3(4), 240. 10.1016/J.WJORL.2017.12.010

25. Smits, C., Theo Goverts, S., & Festen, J. M. (2013). The digits-in-noise test: assessing auditory speech recognition abilities in noise. The Journal of the Acoustical Society of America, 133(3), 1693– 1706. 10.1121/1.4789933

26. Vaerenberg, B., Smits, C., De Ceulaer, G., Zir, E., Harman, S., Jaspers, N., Tam, Y., Dillon, M., Wesarg, T., Martin-Bonniot, D., Gärtner, L., Cozma, S., Kosaner, J., Prentiss, S., Sasidharan, P., Briaire, J. J., Bradley, J., Debruyne, J., Hollow, R., … Govaerts, P. J. (2014). Cochlear implant programming: a global survey on the state of the art. TheScientificWorldJournal, 2014. 10.1155/2014/501738

27. Van Opstal, A. J., & Noordanus, E. (2023). Towards personalized and optimized fitting of cochlear implants. Frontiers in Neuroscience, 17. 10.3389/FNINS.2023.1183126

28. Van Son, R. (1995). A method to quantify the error distribution in confusion matrices. EUROSPEECH.

29. van Wieringen, A., Magits, S., Francart, T., & Wouters, J. (2021). Home-Based Speech Perception Monitoring for Clinical Use With Cochlear Implant Users. Frontiers in Neuroscience, 15. 10.3389/FNINS.2021.773427

30. Walia, A., Shew, M. A., Lefler, S. M., Ortmann, A. J., Durakovic, N., Wick, C. C., Herzog, J. A., & Buchman, C. A. (2023). Factors Affecting Performance in Adults With Cochlear Implants: A Role for Cognition and Residual Cochlear Function. Otology & Neurotology : Official Publication of the American Otological Society, American Neurotology Society [and] European Academy of Otology and Neurotology, 44(10), 988–996. 10.1097/MAO.0000000000004015

31. Warren, S. E., & Atcherson, S. R. (2023). Evaluation of a clinical method for selective electrode deactivation in cochlear implant programming. Frontiers in Human Neuroscience, 17. 10.3389/FNHUM.2023.1157673

32. Wasmann, J. W. A., Huinck, W. J., & Lanting, C. P. (2024). Remote Cochlear Implant Assessments: Validity and Stability in Self-Administered Smartphone-Based Testing. Ear and Hearing, 45(1), 239–249. 10.1097/AUD.0000000000001422

33. Wathour, J., Govaerts, P. J., & Deggouj, N. (2021). Variability of fitting parameters across cochlear implant centres. European Archives of Oto-Rhino-Laryngology : Official Journal of the European Federation of Oto-Rhino-Laryngological Societies (EUFOS) : Affiliated with the German Society for Oto-Rhino-Laryngology - Head and Neck Surgery, 278(12), 4671–4679. 10.1007/S00405-020-06572-W

34. Wathour, J., Govaerts, P. J., Lacroix, E., & Naïma, D. (2023). Effect of a CI Programming Fitting Tool with Artificial Intelligence in Experienced Cochlear Implant Patients. Otology and Neurotology, 44(3), 209–215. 10.1097/MAO.0000000000003810

35. Zeng, F.-G. (2022). Celebrating the one millionth cochlear implant. JASA Express Letters, 2(7). 10.1121/10.0012825

